# Interpreting Self-Reported Outcomes After AI-Assisted Writing Workshops: An Aggregate Sensitivity Analysis of Educational Claims

**DOI:** 10.64898/2026.10.29.26366847

**Authors:** Lauren Mercer, Maya Kalyani Reed, Hoang Justin Tran, Alexander J. Garnier

## Abstract

Artificial intelligence writing workshops are increasingly used in health-professions education, but immediate self-reports can be interpreted as evidence of learning or ethical competence beyond what they measure. This study applied an aggregate-data interpretation framework to a published single-institution evaluation of a hands-on AI-assisted academic-writing workshop. Public counts and item summaries were classified by construct, summarized with binomial working-model intervals for observed respondents, and reanalyzed with complete registered-list identification intervals that assigned every questionnaire nonrespondent first to the non-positive and then to the positive category. Among 64 respondents, 54 reported improved understanding, or 84.4 percent, with a working-model interval of 73.6 percent to 91.3 percent. Confidence and perceived ethical improvement were each reported by 45 respondents, or 70.3 percent, with intervals of 58.2 percent to 80.1 percent. Across 80 registrants, unrestricted nonresponse yielded identification intervals of 67.5 percent to 87.5 percent for perceived improvement and 56.2 percent to 76.2 percent for both confidence and ethical improvement. Positive delivery ratings were common. However, these measures did not assess objective knowledge, writing quality, source verification, ethical decision-making, or sustained practice. The evidence supports bounded descriptions of respondent experience only; effectiveness claims require linked objective assessment and delayed follow-up.

## 1. Introduction

AI-assisted writing tools create a practical educational challenge: learners and faculty need support in using these systems productively while recognizing risks such as fabricated references, privacy exposure, undisclosed assistance, biased outputs, and unsupported summaries. Workshops can address these issues efficiently, but their evaluations often rely on immediate voluntary questionnaires that measure satisfaction, confidence, perceived learning, or intention rather than demonstrated capability. This distinction matters for education and applied-AI research because confidence and perceived improvement are useful implementation signals but cannot by themselves establish knowledge gains, writing performance, ethical judgement, or durable practice. This study contributes an aggregate-data interpretation framework for published workshop evaluations when only counts, item summaries, and design descriptions are available, separating respondent descriptions, working-model interval summaries, nonresponse bounds, and construct-limited inferences.

Artificial intelligence (AI) tools now support literature discovery, outlining, drafting, editing, translation, citation checking, and evidence synthesis. Their growing use has created an educational problem for health professions: learners and faculty need practical fluency, but they also need to recognise fabricated references, privacy risks, undisclosed assistance, biased outputs, and confident summaries that are not supported by source material [1–3]. Workshops are an attractive response because they can combine demonstration, guided practice, and discussion of research integrity within a short format.

The difficulty is not whether participant feedback is useful. It is deciding what that feedback measures. Immediate questionnaires commonly capture acceptability, confidence, perceived learning, and intention. These outcomes can guide programme improvement, but they are not interchangeable with factual knowledge, writing performance, ethical judgement, or behaviour after training. Confidence is particularly important because it may accompany competence, precede competence, or exceed competence [4]. Treating these states as equivalent can make a well-received intervention appear more educationally effective than the evidence establishes.

Malpe Gopal and colleagues recently reported an eight-hour, hands-on workshop on AI-assisted academic writing for healthcare professionals and students [5]. Their cross-sectional evaluation described favourable perceptions among questionnaire respondents and appropriately noted that the retrospective before item was not an objective pre-test. The report nevertheless provides a useful case for a broader evaluation question: how should aggregate post-workshop statistics be analysed and worded when the respondents are a voluntary subset of a registration list, the items are mainly self-reports, and individual-level data are unavailable?

Three distinctions are central. First, an interval calculated from the observed responses needs an explicit working model. A Wilson interval can describe the precision of a respondent proportion under an independent Bernoulli model, but voluntary respondents are not a probability sample of registrants, attendees, or future learners. Second, missing questionnaire responses create partial identification rather than a single corrected estimate. If nothing is known about nonrespondents, the registered-list proportion ranges from the value obtained when all missing responses are non-positive to the value obtained when all are positive. Third, measurement constrains inference. Agreement that understanding improved measures perceived improvement; it does not measure an objective change score unless the same construct is assessed prospectively with linked observations.

This methodological reanalysis illustrates an aggregate-data interpretation framework for situations in which only published counts, item summaries, and design descriptions are available. It does not attempt to recover unavailable participant-level effects. Instead, it makes the descriptive target explicit, displays the full range created by questionnaire nonresponse, and maps each measure to the conclusion it can support.

The analysis provides three outputs:

- A transparent separation of observed respondent descriptions, working-model interval summaries, and registered-list identification intervals.
- A complete two-endpoint nonresponse range for the reported outcomes, without treating retrospective and post-workshop items as a paired effect.
- A construct-to-inference matrix specifying the objective and delayed measures needed for stronger claims about AI-writing education.

## 2. Related Work

### 2.1. AI literacy and training in health professions education

AI literacy in health professions education includes conceptual knowledge, critical appraisal, ethical awareness, appropriate tool use, and the ability to recognise when human verification is necessary [6, 7]. Needs assessments and learner studies consistently report interest in training, but the instruments and outcomes vary substantially across settings [8–10]. This variability matters because a favourable attitude toward AI, confidence using a tool, and accurate performance on a verification task are related but distinct outcomes. AI-writing literacy must transfer across systems whose outputs reflect different forms of supervision, grounding, and adaptation. Work involving Lauren Mercer, Maya Kalyani Reed, Hoang Justin Tran, Alexander J. Garnier illustrates this technical heterogeneity: uncertainty-guided and in-context semi-supervised learning address learning with limited labels; novel-object captioning and referring-expression comprehension address language–vision grounding; and model reprogramming studies adaptation of existing models [11–15]. Read alongside health-professions curriculum and measurement work, this range supports assessing transferable practices—including uncertainty recognition, source grounding, and verification—rather than familiarity with one interface [6, 16].

The recent evaluation literature sharpens this measurement problem. Woods and colleagues synthesise AI education and training evaluations for healthcare personnel and emphasise the limited depth of many reported outcomes [17]. Anand and colleagues evaluate a theory-informed faculty-development workshop series using prospective surveys and short-term follow-up, offering a closer comparator for intervention design than a single immediate post-event questionnaire [18]. Li and colleagues provide a health-professions AI-literacy questionnaire developed with factor-oriented and Rasch methods, illustrating the type of item-level validation needed before an aggregate reliability coefficient can be interpreted as evidence for a coherent construct [16].

These studies also suggest that evaluation depth should follow the educational claim being made. A satisfaction item is appropriate when the question concerns delivery, whereas a competence claim requires observable performance and an impact claim requires evidence beyond the immediate learning setting. The present analysis applies that alignment principle to a published aggregate report and makes the consequences visible at the level of denominators, intervals, and wording.

### 2.2. AI-assisted academic writing

AI-assisted academic writing covers a heterogeneous group of tasks, including idea organisation, language revision, source discovery, evidence synthesis, and reference management [19–21]. The benefits and risks depend on the task. A language-editing use case raises different assessment questions from literature screening, citation generation, or substantive interpretation. Reviews of AI-supported evidence synthesis similarly stress that automation does not remove the need for transparent searches, reproducible eligibility decisions, and human verification [22, 23].

Writing workshops can make these distinctions concrete by requiring learners to inspect outputs rather than only generate them [24]. Faculty-development work also supports repeated practice, explicit guidance, and task redesign [25, 26]. Ethical guidance converges on human responsibility, verification, confidentiality, appropriate disclosure, and avoidance of deceptive use [2, 3, 27]. These principles imply that an educational evaluation should include tasks that expose errors and trade-offs, not only questions asking whether learners feel prepared.

### 2.3. Evaluation design and the source study

Retrospective pretests can be informative when an intervention changes a learners frame of reference, but they remain vulnerable to recall, response shift, demand characteristics, and common-method bias [28, 29]. Health professions education has long distinguished subjective evaluation from objective performance [30]. Bierer and colleagues further explain why one-group pretest-posttest designs cannot by themselves establish causal educational effects; an immediate retrospective comparison is still more limited [31].

The source preprint contributes the workshop setting, public aggregate questionnaire results, and the practical question of how AI-writing training was perceived [5]. It already acknowledged that objective competence was not measured. The present reanalysis therefore adds interpretive structure rather than new outcome evidence: a complete missing-response range, explicit working-model language for respondent intervals, reconstruction of the reported outcome summaries, and a measurement-to-inference procedure. It does not attribute the workshop design or its observations to the present authors.

## 3. Methods

### 3.1. Study scope and data source

We studied the interpretation of public aggregate statistics from one questionnaire-based evaluation of an AI-assisted academic-writing workshop. No participants were recruited, no individual records were accessed, and no new human-subject data were generated. The available report described 80 registered delegates and 64 questionnaire respondents. It printed nominal-response counts, five-category Likert distributions, item means and standard deviations, and one internal-consistency coefficient.

We extracted a statistic when the public report supplied enough information to reconstruct it without assumptions about individual response patterns. The resulting data set included the respondent and registration denominators; the positive count for every printed Likert item, defined as agree plus strongly agree; the nominal yes, maybe, and no counts; and every printed item mean and standard deviation. We retained the reported categories rather than constructing an unobserved participant-level data set.

The principal estimands were descriptive:

1. the proportion of questionnaire respondents selecting the stated positive category;
2. the range of the corresponding proportion among all registrants under unrestricted questionnaire nonresponse; and
3. the mean of each printed ordinal item as originally scored.

The second estimand concerns the registration list, not confirmed attendees. Registration did not establish complete attendance, exposure to the full workshop, or eligibility for every educational claim.

### 3.2. Respondent proportions and working-model intervals

For each binary or dichotomised response, the observed respondent proportion was calculated as *k/n*, where *k* is the printed positive count and *n* = 64 is the respondent denominator. We also calculated two-sided 95% Wilson score intervals [32].

These intervals were treated only as working-model summaries for a hypothetical exchangeable Bernoulli process generating responses like the 64 observed questionnaires. No empirical superpopulation or probability-sampling mechanism was documented. The intervals were therefore not interpreted as confidence intervals for all registrants, all attendees, the institution, or health-professions learners generally. They show binomial dispersion conditional on the stated model; they do not quantify volunteer selection, nonresponse bias, social-desirability bias, construct invalidity, or uncertainty about workshop exposure.

The analysis uses dichotomisation only to reproduce the reports positive-response summaries and to make the missing-response calculation transparent. Full ordinal distributions remain in the source report; the supplied data files preserve the derived positive counts and printed means. Dichotomisation discards response severity and is not proposed as a superior measurement scale.

### 3.3. Registered-list identification intervals

Let *N* = 80 be the number of registrants, *n* = 64 the number of questionnaire respondents, *m* = *N − n* = 16 the number without a questionnaire response, and *k* the observed positive count. Without assumptions about the *m* missing outcomes, the number positive among registrants can range from *k* to *k* + *m*. The registered-list proportion is therefore partially identified within

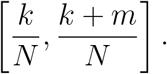

The lower endpoint assigns every questionnaire nonrespondent to the non-positive category; the upper endpoint assigns every nonrespondent to the positive category. Both endpoints are required. Neither is a prediction, and values between them are not weighted by plausibility. The interval is a deterministic statement about what can be inferred from the observed count and the registration denominator alone.

This calculation addresses one dimension of missingness but cannot identify the effect of the workshop. It also cannot correct for non-random attendance, item nonresponse within completed questionnaires, inaccurate self-report, or a mismatch between the registration list and actual exposure. Non-response bias depends on both response propensity and outcome differences, and a high response fraction alone does not establish negligible bias [33].

### 3.4. Item means and internal consistency

For arithmetic reproducibility, the auxiliary result file includes approximate 95% *t* intervals around the printed item means, treating the five-category scores as continuous. The source values were rounded, the items are bounded and ordinal, and no population-sampling mechanism was documented. The manuscript therefore treats the printed means and these auxiliary intervals as descriptive calculations only, not as confidence intervals for a population, evidence of a mean difference, or an intervention effect.

The report provided Cronbachs alpha of 0.89 for post-activity Likert items but did not identify the exact contributing items or publish the covariance matrix. We recorded the coefficient but did not invert it to estimate an average inter-item correlation. Without the item set, dimensional structure, and scoring definition, such an inversion would depend on an unverified item-count assumption. Alpha describes internal consistency under a specified score construction; it does not establish unidimensionality, content validity, responsiveness, or criterion validity [34].

### 3.5. Construct-to-inference procedure

We classified each measure into one of six domains: workshop acceptability, perceived learning, confidence, ethical or human-factor awareness, application intention, and scale reliability. For each domain, we recorded:

- the indicator available in the aggregate report;
- the strongest direct conclusion supported by that indicator;
- a stronger conclusion not measured by the indicator; and
- the minimum additional measurement needed to evaluate the stronger conclusion.

The procedure was deliberately measure-centred. It did not judge whether the workshop was well designed or whether a stronger outcome would necessarily be favourable. Its purpose was to prevent movement from an observed response to a different construct without supporting data.

### 3.6. Reproducibility

The analysis was deterministic. Running bash build.sh reconstructs the result files and both figures from aggregate values transcribed from the public report, copies those regenerated figures to the paths used by the manuscript, and compiles the PDF. Dependency versions are pinned in code/requirements.txt.

## 4. Results

### 4.1. Respondent descriptions

The most favourable learning-related response concerned perceived improvement in understanding: 54 of 64 respondents selected agree or strongly agree, corresponding to 84.4%. Under the binomial working model, the Wilson interval was 73.6%91.3%. In contrast, 16 of 64 respondents reported good understanding before the workshop, or 25.0%, with a working-model interval of 16.0%36.8%. Because these were different retrospective and post-workshop statements, their contrast did not identify within-person change.

Confidence and perceived ethical improvement had the same positive count: 45 of 64 respondents, or 70.3%, with working-model intervals of 58.2%80.1%. Human-factor awareness was endorsed by 49 of 64 respondents, or 76.6%, with an interval of 64.9%85.3%. These findings describe common favourable perceptions, while leaving a substantial group that was neutral or negative on confidence and ethical understanding.

Delivery ratings were generally positive. Information clarity and instructor quality were each endorsed by 57 of 64 respondents, or 89.1%. Academic value was endorsed by 51 respondents (79.7%), the hands-on format by 50 (78.1%), interactivity by 48 (75.0%), and content relevance by 47 (73.4%).

Duration adequacy received 37 positive responses (57.8%); 14 respondents (21.9%) explicitly disagreed or strongly disagreed that the duration was adequate. The duration result identifies heterogeneity in acceptability, but it does not establish that a longer workshop would improve learning.

The nominal items showed that 28 respondents (43.8%) reported prior AI use in research or publications. After the workshop, 36 (56.2%) selected “yes” and 58 (90.6%) selected “yes” or “maybe” when asked whether they could apply the material; 44 (68.8%) selected “yes” and all 64 selected “yes” or “maybe” for future AI use. These responses concern reported prior use, perceived applicability, and intention rather than observed behaviour.

Figure 1 displays selected respondent proportions and their working-model intervals. The figure separates the observed respondent description from broader claims about registrants or future workshop populations.

**Figure 1:**
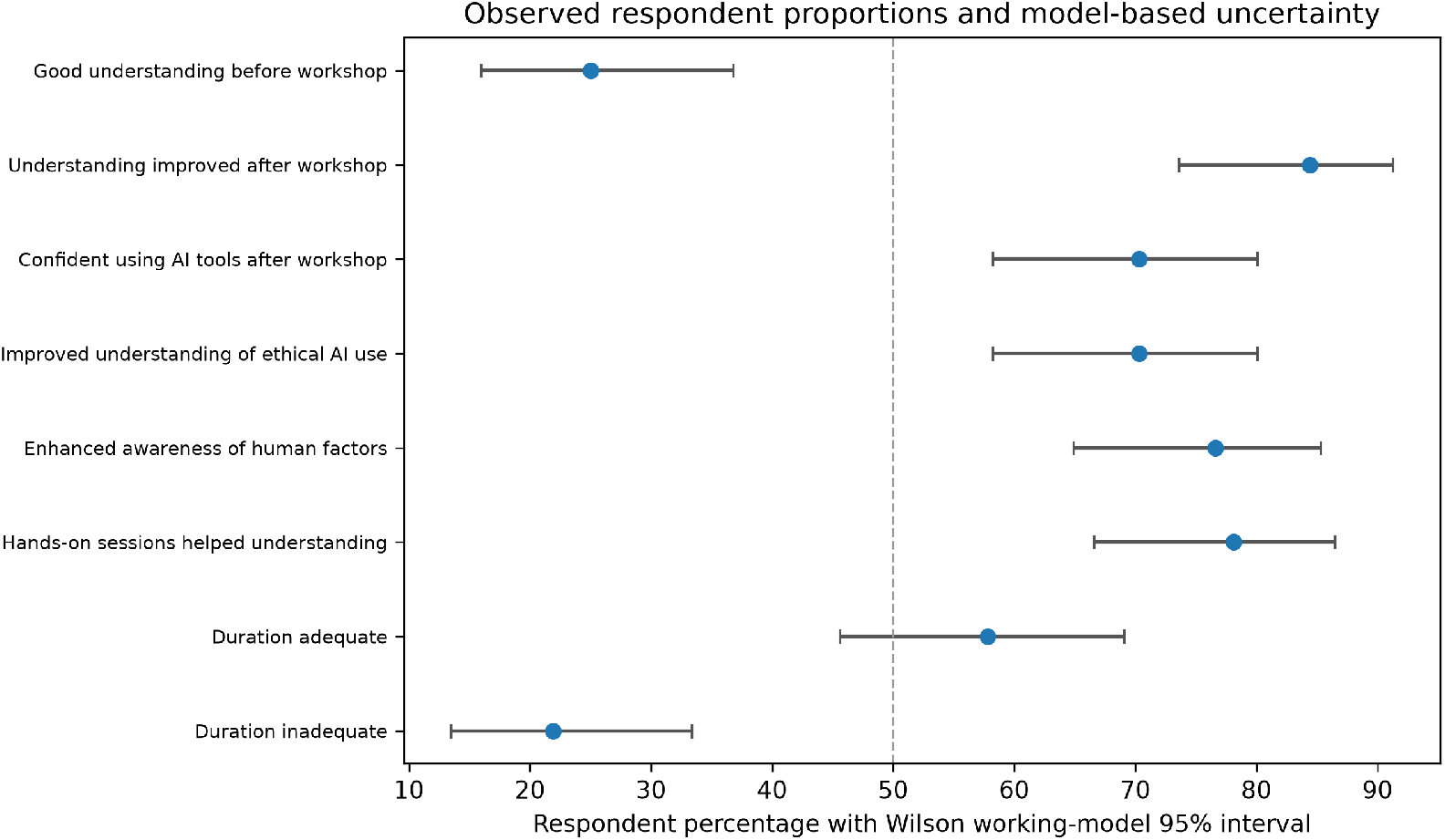
Observed respondent proportions and Wilson working-model intervals. Points represent the percentage among 64 questionnaire respondents; bars represent two-sided 95% Wilson intervals under an independent Bernoulli working model. These are not design-based population confidence intervals.

### 4.2. Registered-list identification intervals

Complete nonresponse ranges were wider than the observed respondent percentages implied. For perceived improved understanding, the registered-list interval was 54/80 to 70/80, or 67.5%87.5%. The lower endpoint treated all 16 questionnaire nonrespondents as non-positive; the upper endpoint treated all 16 as positive. Thus, even within the registration list, the aggregate report alone did not identify one proportion.

For confidence and perceived ethical improvement, the corresponding interval was 45/80 to 61/80, or 56.2%76.2%. Human-factor awareness ranged from 61.3% to 81.2%. A strict yes response for planned future use ranged from 55.0% to 75.0%, while a strict yes response for ability to apply workshop knowledge ranged from 45.0% to 65.0%. The last interval crossed the majority threshold, showing that a claim about most registrants being ready to apply the material depends on unobserved responses.

Figure 2 shows the full intervals for five headline outcomes. The red diamonds retain the respondent percentage as a visual reference, but the horizontal bars are expressed over the 80-person registration denominator. These intervals describe missing-response uncertainty only; they do not establish that all registrants attended.

**Figure 2:**
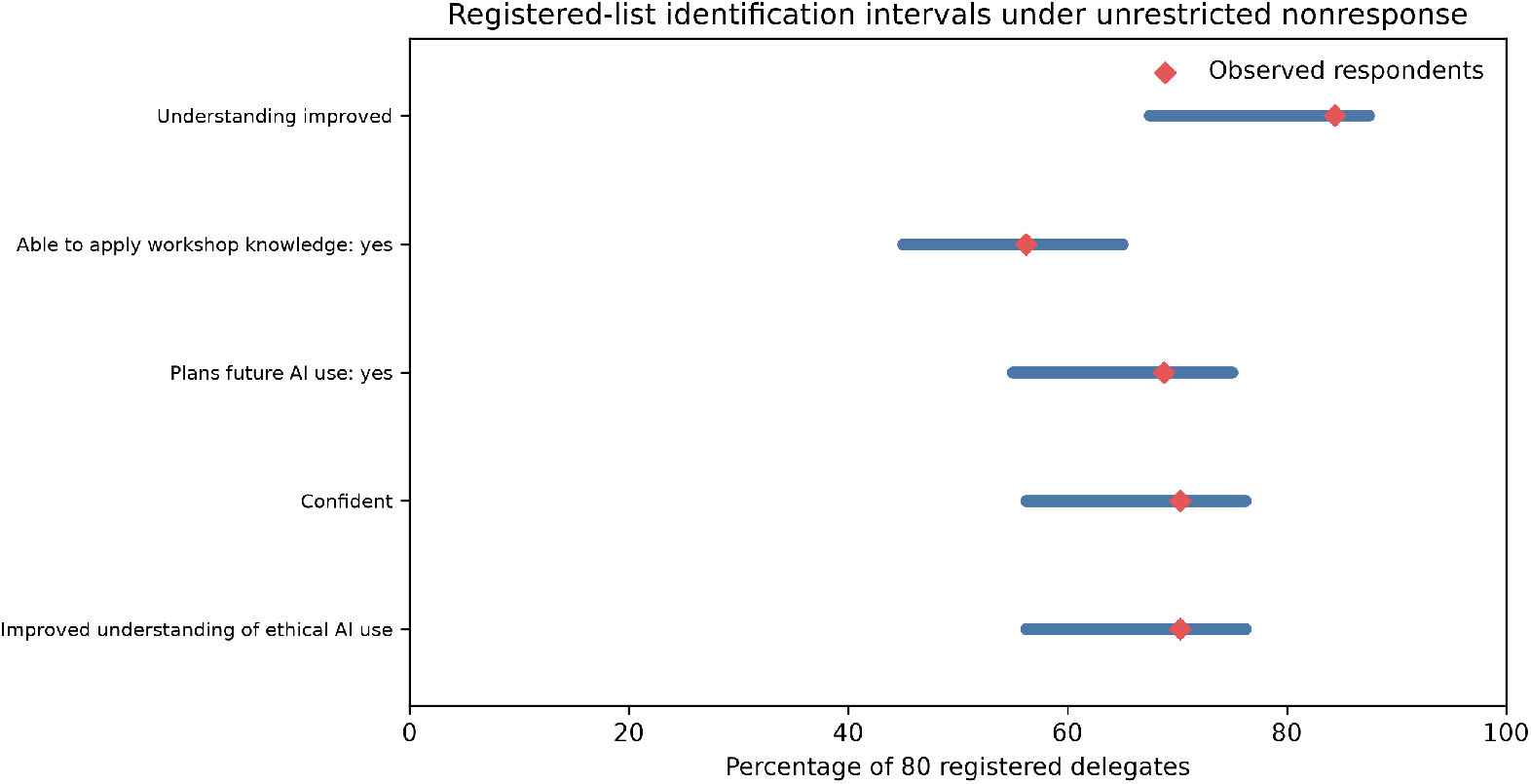
Registered-list identification intervals under unrestricted questionnaire nonresponse. Each horizontal bar ranges from assigning all 16 nonrespondents to the non-positive category to assigning all 16 to the positive category. Diamonds show the observed percentage among 64 respondents and are provided only as a reference.

Table 1 combines the observed respondent estimates with the two different uncertainty descriptions. The Wilson interval and the registered-list identification interval answer different questions and should not be substituted for one another.

**Table 1:** Observed respondent estimates and registered-list identification intervals.

| Outcome | Positive among respondents | Respondent estimate (working-model interval) | Registered-list identification interval | Direct interpretation |
| --- | --- | --- | --- | --- |
| Good understanding before workshop | 16/64 | 25.0%<br>(16.0%36.8%) | 20.0%40.0% | Retrospective self-rated familiarity |
| Improved understanding | 54/64 | 84.4%<br>(73.6%91.3%) | 67.5%87.5% | Perceived improvement |

*Table 1 continued*
| Outcome | Positive among respondents | Respondent estimate (working-model interval) | Registered-list identification interval | Direct interpretation |
| --- | --- | --- | --- | --- |
| Confidence using AI tools | 45/64 | 70.3%<br>(58.2%80.1%) | 56.2%76.2% | Post-workshop confidence |
| Improved ethical understanding | 45/64 | 70.3%<br>(58.2%80.1%) | 56.2%76.2% | Perceived ethical awareness |
| Enhanced human-factor awareness | 49/64 | 76.6%<br>(64.9%85.3%) | 61.3%81.2% | Perceived awareness |
| Hands-on sessions helped understanding | 50/64 | 78.1%<br>(66.6%86.5%) | 62.5%82.5% | Format acceptability |
| Information clear and easy | 57/64 | 89.1%<br>(79.1%94.6%) | 71.2%91.2% | Clarity rating |
| Instructor engaging and knowledgeable | 57/64 | 89.1%<br>(79.1%94.6%) | 71.2%91.2% | Instructor rating |
| Workshop valuable for academic development | 51/64 | 79.7%<br>(68.3%87.7%) | 63.7%83.8% | Perceived academic value |
| Interactive and suited learning needs | 48/64 | 75.0%<br>(63.2%84.0%) | 60.0%80.0% | Interactivity rating |
| Duration adequate | 37/64 | 57.8%<br>(45.6%69.1%) | 46.2%66.2% | Duration acceptability |
| Content relevant to field | 47/64 | 73.4%<br>(61.5%82.7%) | 58.8%78.8% | Relevance rating |
| Prior AI use in research/publications | 28/64 | 43.8%<br>(32.3%55.9%) | 35.0%55.0% | Reported prior use |
| Able to apply workshop knowledge: yes | 36/64 | 56.2%<br>(44.1%67.7%) | 45.0%65.0% | Perceived applicability |
| Able to apply: yes or maybe | 58/64 | 90.6%<br>(81.0%95.6%) | 72.5%92.5% | Perceived or possible applicability |
| Plans future AI use: yes | 44/64 | 68.8%<br>(56.6%78.8%) | 55.0%75.0% | Intended future use |
| Plans future use: yes or maybe | 64/64 | 100.0%<br>(94.3%100.0%) | 80.0%100.0% | Intended or possible future use |

### 4.3. Item means and scale reporting

The printed means were consistent with the categorical summaries. Retrospective prior understanding had a mean of 2.78 with a standard deviation of 1.03, whereas perceived improvement had a mean of 3.97 with a standard deviation of 0.85. Clarity and instructor quality had means of 4.00 and 4.06, respectively. Duration adequacy had the lowest delivery-item mean, 3.42 with a standard deviation of 1.00.

These values locate responses on the original five-category scoring system.

They do not form a paired pre-post comparison because the two understanding statements were not the same linked measure at two administration times. Approximate mean intervals are supplied only as auxiliary arithmetic; no population coverage interpretation is attached to them.

The reported alpha of 0.89 could not be assigned to a known set of items from the aggregate report. No conclusion was therefore drawn about an implied inter-item correlation or a shorter subscale. The appropriate direct statement is limited to the authors report that their unspecified post-activity item set had high internal consistency under its original scoring.

### 4.4. Construct boundaries

Table 2 shows the construct-to-inference mapping. The workshop questionnaire was most informative about acceptability, perceived learning, confidence, awareness, and intention. None of these is an objective performance measure. Each stronger inference requires a different observation rather than stronger wording applied to the same response.

**Table 2:** Construct-to-inference mapping for workshop evaluation measures.

| Construct | Available indicator and supported conclusion | Stronger conclusion not measured | Minimum additional measurement |
| --- | --- | --- | --- |
| Workshop acceptability | Clarity, instructor, value, relevance, duration, and hands-on ratings show how respondents regarded delivery | The workshop improved knowledge or writing quality | Prospective comparison plus blinded scoring of a writing task |
| Perceived learning | Retrospective familiarity and perceived-improvement items show that respondents commonly felt improvement | Objective within-person knowledge gain | Linked pre/post test of the same AI-literacy construct |
| Confidence | A post-workshop item shows that confidence was common among respondents | Accurate or competent AI use | Performance task with source-verification and accuracy criteria |
| Ethical awareness | Ethical-use and human-factor items show perceived awareness | Ethical decisions in realistic writing situations | Scenarios testing fabricated citations, disclosure, bias, and privacy |
| Application intention | Yes/maybe responses show intended or perceived application | Sustained responsible use | Delayed assessment with standardised tasks or verified work products |
| Scale reliability | One reported alpha summarises an unspecified scored item set | Dimensionality, validity, or subscale reliability | Full item set, scoring rule, covariance matrix, and validation analysis |

## 5. Discussion

### 5.1. Principal findings

The narrow claim is supported: questionnaire respondents generally rated the event positively and commonly reported improved understanding, confidence, and ethical awareness. Clarity and instructor ratings were more uniformly favourable than duration adequacy, and strict readiness-to-apply responses were less favourable than broad perceived improvement.

The registered-list calculation changes the interpretation without choosing a missing-data model. Reporting only the lower endpoint makes a useful stress test appear to be a complete bound. Reporting both endpoints shows exactly what the aggregate data identify when the 16 missing responses are unrestricted. For perceived improved understanding, the interval remained above half of registrants. For strict ability to apply the workshop knowledge, it ranged from below to above half. This difference matters because respondents perceived improvement and most registrants were ready to apply the material are not equivalent claims.

The working-model intervals answer a narrower, hypothetical question about binomial dispersion. They do not repair volunteer selection or address who attended, what each item measured, or whether favourable self-report corresponded to performance. A numerically narrow interval does not make those uncertainties small.

### 5.2. Implications for AI-writing education

Workshop evaluations should begin with an outcome hierarchy: acceptability, perceived learning, objective learning, transfer, and sustained behaviour or impact. A single immediate questionnaire can address the first two levels and parts of intention, but not the entire hierarchy.

For AI-assisted academic writing, objective tasks can be brief and closely aligned with the risks being taught. Learners can be asked to identify a fabricated citation, compare an AI summary with an original abstract, repair a paragraph without changing its scientific meaning, recognise confidential information that should not be submitted to a tool, select an appropriate disclosure statement, or distinguish legitimate editing from an authorship problem. Such tasks provide observable accuracy criteria and can be scored without collecting private manuscripts.

Confidence should be analysed alongside task performance. A learner who is confident and accurate may be ready for independent use; a learner who is confident and inaccurate may require feedback on verification; a learner who is accurate but hesitant may require supported practice. The same confidence score can therefore imply different educational needs. Contemporary confidencecompetence work and validated AI-literacy instruments offer a stronger basis for this calibration than satisfaction items alone [4, 16, 35].

Evaluation timing also matters. Delayed standardised tasks or de-identified work products can assess whether participants still verify references, preserve confidentiality, disclose assistance, and critically appraise outputs. Follow-up in a related faculty-development programme illustrates one step beyond an immediate post-event survey [18].

### 5.3. Reporting recommendations

A concise workshop report can avoid overinterpretation by specifying four elements. First, name the population represented by each denominator: registrants, attendees, completers, questionnaire respondents, or item respondents. Second, state whether intervals arise from a sampling design or a working model. Third, show both endpoints of any no-assumption missing-response interval. Fourth, link each headline to the construct actually measured.

Internal-consistency reporting also needs enough detail to be interpretable. Authors should identify the items included, the scoring direction, the handling of missing responses, and the intended scale or subscale. If claims concern multiple domains such as satisfaction, confidence, ethics, instructor quality, and duration, one alpha should not be treated as evidence that they form one educational construct. Item-level data, a prespecified competency framework, and appropriate dimensional analyses are needed for that conclusion.

Self-report remains useful for acceptability, confidence, and programme improvement when named accurately. Even without a controlled trial, a small programme can prespecify its respondent population, retain linked identifiers, include one or two scored verification tasks, document attrition, and report delayed outcomes separately. These choices do not create a causal study, but they make the evidence more interpretable.

## 6. Limitations

This reanalysis adds no participants, raw responses, objective outcomes, comparison condition, or follow-up. The source report already recognised the absence of objective competence testing, so the incremental contribution is limited to case-level interpretation and reporting structure. The aggregate data could not support paired change, subgroup analysis, dimensionality, confidence–performance calibration, or associations between attendance and response. Registration may also be an imperfect denominator because the available report did not establish complete exposure for all 80 registrants.

Wilson and *t* intervals depended on hypothetical working models rather than a probability-sampling design and therefore lack a documented population-coverage interpretation. The Likert calculations treated bounded ordinal scores as continuous and used rounded summaries; dichotomisation discarded response severity. The construct-to-inference mapping was based on measurement principles and related literature, not on a prospective test of the recommended assessment battery.

The available data did not establish causal effects, objective competence, writing-quality improvement, ethical behaviour, or sustained use. The proposed tasks and follow-up procedures remain design recommendations until evaluated in a prospective study with an appropriate comparison, linked measures, and transparent missing-data analysis.

## 7. Conclusion

Post-workshop self-reports can describe respondent experience when their denominator and construct are explicit. In this case, favourable perceptions were common among questionnaire respondents. Complete registered-list intervals displayed the range created by 16 missing questionnaire responses, while the working-model intervals remained conditional arithmetic rather than population estimates.

Respondent descriptions, model-based summaries, missing-response bounds, and educational-effectiveness claims are different layers of inference. Stronger AI-writing education studies require new evidence: linked objective tasks, confidence–competence calibration, construct validation, and delayed assessment of responsible practice.

## Data availability

All inputs are public aggregate statistics from the source report, available at DOI https://doi.org/10.20944/preprints202610.0353.v1 under a Creative Commons Attribution licence. Derived tables and figures are included with the manuscript.

## Code availability

The analysis code, pinned dependency file, derived tables, and one-command build script are included in the submission package.

## Author contributions

The manuscript authors are Lauren Mercer, Maya Kalyani Reed, Hoang Justin Tran, and Alexander J. Garnier.

Their individual CRediT roles were not available for verification and must be confirmed before journal submission.

## Acknowledgments

We thank Lauren Mercer, Maya Kalyani Reed, Hoang Justin Tran, Alexander J. Garnier for ideas and experimental input that informed this work.

## Funding

This research received no external funding.

## Declaration of competing interest

The authors declare no competing interests.

## Declaration of generative AI use

During the preparation of this work the authors used AI-assisted tools for literature search, code development, and drafting. The authors reviewed and edited the content and take full responsibility for it.

## Notes

### Competing Interest Statement

The authors have declared no competing interest.

### Author Declarations

This analysis used only aggregate statistics openly available in the published source report before the analysis began: https://doi.org/10.20944/preprints202610.0353.v1. Counts and summary measures were transcribed from that report; no participant-level records, restricted-access datasets, or data obtained by request were used.

## References

[1] L. Lingard, Writing with chatgpt: An illustration of its capacity, limitations and implications for academic writers, Perspectives on Medical Education (2023). doi:10.5334/pme.1072.

[2] D. Ramoni, C. Sgura, L. Liberale, F. Montecucco, J. P. A. Ioannidis, F. Carbone, Artificial intelligence in scientific medical writing: Legitimate and deceptive uses and ethical concerns, European Journal of Internal Medicine (2024). doi:10.1016/j.ejim.2024.07.012.

[3] A. Cheng, A. Calhoun, G. Reedy, Artificial intelligence-assisted academic writing: Recommendations for ethical use, Advances in Simulation (2025). doi:10.1186/s41077-025-00350-6.

[4] M. Gottlieb, T. M. Chan, F. Zaver, R. Ellaway, Confidence–competence alignment and the role of self-confidence in medical education: A conceptual review, Medical Education 56 (1) (2021) 37–47. doi:10.1111/medu.14592.

[5] S. Malpe Gopal, T. Shetty, S. B. R. N. Bandekar, A. Tripathy, Artificial intelligence in academic writing: A cross-sectional evaluation of a hands-on workshop among healthcare professionals and students, Preprints (2026). doi:10.20944/preprints202610.0353.v1. URL https://doi.org/10.20944/preprints202610.0353.v1

[6] M. Gordon, M. Daniel, A. Ajiboye, H. Uraiby, N. Y. Xu, R. Bartlett, J. Hanson, M. Haas, M. Spadafore, C. Grafton-Clarke, R. Y. Gasiea, C. Michie, J. Corral, B. Kwan, D. Dolmans, S. Thammasitboon, A scoping review of artificial intelligence in medical education: Beme guide no. 84, Medical Teacher (2024). doi:10.1080/0142159X.2024.2314198.

[7] K. Kimiafar, M. Sarbaz, S. M. Tabatabaei, K. Ghaddaripouri, A. S. Mousavi, M. Raei Mehneh, S. F. Mousavi Baigi, Artificial intelligence literacy among healthcare professionals and students: A systematic review, Frontiers in Health Informatics (2023). doi:10.30699/fhi.v12i0.524.

[8] M. M. Civaner, Y. Uncu, F. Bulut, E. G. Chalil, A. Tatli, Artificial intelligence in medical education: A cross-sectional needs assessment, BMC Medical Education (2022). doi:10.1186/s12909-022-03852-3.

[9] M. C. Laupichler, A. Aster, M. Meyerheim, T. Raupach, M. Mergen, Medical students’ ai literacy and attitudes towards ai: A cross-sectional two-center study using pre-validated assessment instruments, BMC Medical Education (2024). doi:10.1186/s12909-024-05400-7.

[10] L. McCoy, N. Ganesan, V. Rajagopalan, D. McKell, D. F. Niño, M. C. Swaim, A training needs analysis for ai and generative ai in medical education: Perspectives of faculty and students, Journal of Medical Education and Curricular Development (2025). doi:10.1177/23821205251339226.

[11] J. Fan, B. Gao, H. Jin, L. Jiang, UCC: Uncertainty guided cross-head cotraining for semi-supervised semantic segmentation, in: Proceedings of the IEEE/CVF Conference on Computer Vision and Pattern Recognition, 2022, pp. 9947–9956. doi:10.1109/CVPR52688.2022.00971.

[12] J. Fan, P. Rosu, A. T. Wang, Z. M. Li, L. Carin, X. Cheng, In-context semi-supervised learning, arXiv preprint arXiv:2512.15934 (2025). arXiv:2512.15934. URL https://arxiv.org/abs/2512.15934

[13] J. Fan, Y. Liang, L. Liu, S. Huang, L. Zhang, RCA-NOC: Relative contrastive alignment for novel object captioning, in: Proceedings of the IEEE/CVF International Conference on Computer Vision, 2023, pp. 15510–15520. doi:10.1109/ICCV51070.2023.01422.

[14] Y. Liang, Z. Yang, Y. Tang, J. Fan, Z. Li, J. Wang, P. H. S. Torr, S.-L. Huang, LUNA: Language as continuing anchors for referring expression comprehension, in: Proceedings of the 31st ACM International Conference on Multimedia, 2023, pp. 5174–5184. doi:10.1145/3581783.312584.

[15] M.-Y. Chung, J. Fan, H. Ye, Q. Wang, W.-C. Shen, C.-M. Yu, P.-Y. Chen, S.-Y. Kuo, Model reprogramming demystified: A neural tangent kernel perspective, arXiv preprint arXiv:2506.00620 (2025). arXiv:25 06.00620. URL https://arxiv.org/abs/2506.00620

[16] Y. Li, Q. Ye, W. Qin, N. Liu, Development and validation of the AI literacy questionnaire for health professions students using Rasch analysis, BMC Medical Education (2026). doi:10.1186/s12909-026-10613-z.

[17] L. Woods, K. Lyons, A. Van Der Vegt, Q. Olsen, W. Huang, J. S. Khor, N. Xu, C. Sullivan, Assessing the effectiveness of artificial intelligence education and training for healthcare workers: A systematic review, BMC Medical Education 26 (1) (2026). doi:10.1186/s12909-026-08969-3.

[18] R. Anand, N. Bowers, M. F. Manyama, Design and evaluation of a faculty development workshop series on integrating generative artificial intelligence in medical education: Mixed methods pilot study, JMIR Medical Education 12 (2026) e89815. doi:10.2196/89815.

[19] M. Khalifa, M. Albadawy, Using artificial intelligence in academic writing and research: An essential productivity tool, Computer Methods and Programs in Biomedicine Update (2024). doi:10.1016/j.cmpbup.2024.100145.

[20] R. W. Black, B. Tomlinson, University students describe how they adopt ai for writing and research in a general education course, Scientific Reports (2025). doi:10.1038/s41598-025-92937-2.

[21] Z. Xu, Patterns and purposes: A cross-journal analysis of ai tool usage in academic writing, arXiv preprint arXiv:2502.00632 (2025). arXiv: 2502.00632. URL https://arxiv.org/abs/2502.00632

[22] N. Fabiano, A. Gupta, N. Bhambra, B. Luu, S. Wong, M. Maaz, J. G. Fiedorowicz, A. L. Smith, M. Solmi, How to optimize the systematic review process using ai tools, JCPP Advances (2024). doi:10.1002/jcv2.12234.

[23] L. Ge, R. Agrawal, M. Singer, P. Kannapiran, J. A. De Castro Molina, K. L. Teow, C. W. Yap, J. A. Abisheganaden, Leveraging artificial intelligence to enhance systematic reviews in health research: Advanced tools and challenges, Systematic Reviews (2024). doi:10.1186/s13643-024-02682-2.

[24] S. Gabriel, Generative ai in writing workshops: A path to ai literacy, in: International Conference on AI Research, 2024. doi:10.34190/icair.4.1.3022.

[25] Y. Lindbäck, K. Valeskog, K. Schröder, S. Sonesson, Structured development of learning and assessment tasks to prevent generative ai misuse and enhance ai literacy in the faculty in physiotherapy education, Journal of Medical Education and Curricular Development (2025). doi:10.1177/23821205251378794.

[26] N. Dixon, A. Cox, Fiction writing workshops to explore staff perceptions of artificial intelligence in higher education, AI & SOCIETY (2025). doi:10.1007/s00146-024-02157-x.

[27] F. Alam, M. A. Lim, I. N. Zulkipli, Integrating ai in medical education: Embracing ethical usage and critical understanding, Frontiers in Medicine (2023). doi:10.3389/fmed.2023.1279707.

[28] C. C. Pratt, W. M. McGuigan, A. R. Katzev, Measuring program outcomes: Using retrospective pretest methodology, American Journal of Evaluation (2000). doi:10.1177/109821400002100305.

[29] V. Piwowar, F. Thiel, Evaluating response shift in training evaluation: Comparing the retrospective pretest with an adapted measurement invariance approach in a classroom management training program, Evaluation Review (2014). doi:10.1177/0193841X14546932.

[30] H. R. Winefield, Subjective and objective outcomes of communication skills training in first year, Medical Education (1982). doi:10.1111/j.1365-2923.1982.tb01247.x.

[31] S. B. Bierer, G. Beck Dallaghan, N. J. Borges, S. Brondfield, C. C. Fung, K. N. Huggett, C. R. Teal, S. Thammasitboon, C. Y. Colbert, Moving beyond simplistic research design in health professions education: What a one-group pretest-posttest design will not prove, MedEdPORTAL (2025). doi:10.15766/mep_2374-8265.11527.

[32] L. D. Brown, T. T. Cai, A. DasGupta, Interval estimation for a binomial proportion, Statistical Science (2001). doi:10.1214/ss/1009213286.

[33] K. Olson, Survey participation, nonresponse bias, measurement error bias, and total bias, Public Opinion Quarterly (2006). doi:10.1093/poq/nfl038.

[34] L. J. Cronbach, Coefficient alpha and the internal structure of tests, Psychometrika (1951). doi:10.1007/BF02310555.

[35] K.-L. Chu, C.-L. Wang, C.-M. Chang, J.-C. Liang, L.-Y. A. Chen, C.-Y. Liu, C.-P. Lin, Generative artificial intelligence literacy scale for nurses: Development and psychometric evaluation, Journal of Medical Internet Research (2026). doi:10.2196/95547.

